# Collecting data on HIV-related mortality during household surveys: a randomized validation study in Malawi

**DOI:** 10.1101/2020.06.01.20118810

**Authors:** Abena Amoah, Sarah Brumfield, Amelia C. Crampin, Albert N. Dube, Stéphane Helleringer, Estelle McLean, Georges Reniers

## Abstract

**Background:** In many African countries, there are limited representative data on HIV/AIDS mortality. We tested whether such data could be collected during household surveys periodically conducted in most African countries.

**Methods:** We added HIV questions to the module on adult and maternal mortality used in Demographic and Health Surveys. We conducted a validation study of the data generated by these questions in northern Malawi. We randomly assigned men and women aged 15–59 years old to a face-to-face interview or audio computer-assisted self-interviewing (ACASI). We compared survey reports of adult deaths to prospective data on mortality and HIV collected by the Karonga Health and Demographic Surveillance Site. We calculated the sensitivity and specificity of survey data in recording the HIV status of deceased siblings of respondents. We adjusted for partial verification bias using multiple imputations.

**Results:** We interviewed 535 participants, who reported 885 deaths at ages 15 and older. The added HIV questions yielded largely complete data on the HIV status of respondents’ deceased siblings, particularly those who died recently. The adjusted sensitivity of survey data on HIV status of the deceased was high in both study groups (0.78–0.82). There were few false positive reports of the HIV status of deceased siblings (specificity = 0.96–0.98). ACASI did not improve the accuracy of survey data, but it required more time to collect mortality reports. Asking the HIV questions only took 0.4 minute (≈25 seconds) per deceased sibling in face-to-face interviews.

**Conclusions:** Adding HIV questions to mortality questionnaires used in household surveys yields accurate data on the HIV status of deceased adults. These new data could aid in tracking progress towards global HIV elimination targets.

MEIRU – LSHTM – JHU Partnership for improving adult mortality data^1,2^

## INTRODUCTION

Reaching “zero AIDS-related deaths” by 2030 is a key goal of HIV prevention and treatment programs worldwide.^1^ In most African countries, the data required to track progress towards this objective are limited.^2^ Reliable data on HIV/AIDS mortality are only available for select populations. Clinical cohorts^3,4^ measure mortality among patients on antiretroviral therapy (ART), but they rarely document the survival of persons with HIV (PHIV) who are not on ART and they exclude HIV-negative persons. Health and demographic surveillance systems (HDSS) collect population-based data on causes of death,^5,6^ but they often cover small, rural populations.

In the absence of representative mortality data, national estimates of HIV/AIDS deaths in most African countries are inferred primarily from data on HIV prevalence and ART coverage.^7,8^ Several statistical and epidemiological models exist for this purpose.^8,9^ They can yield discrepant estimates of the number of lives saved by HIV programs.^10^ To better calibrate the models tracking progress towards zero AIDS-related deaths in African countries, we tested whether data on HIV/AIDS mortality could be collected during household surveys.

Household surveys such as the demographic and health surveys (DHS) or the multiple indicator cluster surveys (MICS) are conducted periodically in most African countries,^11,12^ where they constitute the main source of representative data on all-cause mortality.^13^ They ask respondents to report the survival of close relatives.^14^ They include questions about pregnancy-related deaths^14^ and injuries^15^, but they do not inquire about HIV/AIDS mortality because of concerns that respondents may not know whether their relatives died of HIV-related causes. These concerns might now have receded, due to a) the scale-up of HIV testing,^16^ b) increased disclosure of HIV status to family members^17,18^ and/or c) conversations in social networks to determine if a death can be attributed to HIV/AIDS.^19^ Survey respondents may also be more willing to disclose such information following declines in HIV stigma.^20,21^

We inserted questions about the HIV status of a respondent’s deceased relatives into the standard DHS module on adult/maternal mortality. These HIV questions do not allow classifying causes of deaths using codes from the International Classification of Diseases. However, by comparing HIV prevalence among survivors and deceased relatives, they allow measuring excess mortality associated with HIV (appendix A1). We conducted a validation study of the data generated by these questions in Northern Malawi.

## Data and Methods

*Data sources*

To ascertain the HIV status of deceased relatives, we adapted existing questions about a respondent’s own HIV status.^22^ We asked respondents if their relative had ever been tested, and if so what were the results of their most recent test. To limit missing data, we asked respondents to assess the likelihood that their relative was infected with HIV. Response categories ranged from “highly likely” to “highly unlikely”. This question was not applicable if the respondent’s relative had received positive test results.

We included the HIV questions in siblings’ survival histories, i.e., a questionnaire wherein respondents list all their maternal siblings, state whether they are alive, and report the current age of live siblings, or the age at death and time since death of deceased siblings.^23^ We asked HIV questions for all deaths reported at ages 15 years and older. We classified deceased siblings as PHIV when respondents stated that the deceased had received positive test results or was “likely” or “highly likely” to have been infected with HIV. The HIV status of the deceased was coded as “missing” if the respondent did not know the likelihood that their sibling was infected with HIV. All other deaths were deaths of HIV-negative persons.

We used data collected prospectively in Karonga district as reference against which we evaluated survey data on HIV-related mortality. This an area of Malawi where the local economy is dominated by subsistence farming, fishing and small-scale trading. Since 2002, a HDSS covers a predominantly rural population of ≈40,000 people in the southern part of Karonga district.^24,25^ After a baseline census, key informants from HDSS villages continuously record births, deaths and migrations among the HDSS population. Each HDSS resident is attributed a unique identification (ID) number. Parental IDs are added to individual HDSS records. This allows listing the known siblings of most HDSS residents, by looking up records that share parental IDs.

Extensive HIV data are available in Karonga. Prior to the HDSS, studies of leprosy and tuberculosis have included HIV testing.^26,27^ HDSS residents were then offered HIV testing repeatedly between 2005 and 2011.These serosurveys achieved participation rates > 70%, and many participants learned their HIV status.^28^ HIV-related data (including HIV test results) from registers at health facilities serving HDSS communities have been linked to HDSS records.^29–31^ Ancillary studies have asked selected HDSS residents to self-report the result and date of recent HIV tests.^32,33^ The HDSS also elicits the HIV status of deceased residents during post-mortem verbal autopsies, but we did not consider these data due to limited reliability.^34^

We constructed a reference classification of adult deaths by HIV status of the deceased, based solely on pre-mortem data from prospective datasets. Deaths of PHIV were deaths for which a clinical record or self-report of an HIV-positive test was available. Deaths of persons who were HIV-negative were deaths for which there was no evidence of a positive test, but a clinical record or self-report of an HIV-negative test was available within five years of the death.^35,36^ Deaths without evidence of a positive or negative HIV test had missing reference data on HIV status.

### Data collection and linkages

We selected study participants at random among HDSS residents aged 15 to 59 years old. We oversampled residents with one or more adult death(s) among known siblings. We oversampled at a higher rate the residents whose deceased sibling(s) was(ere) classified as PHIV by reference data. We selected at most one participant per family. We recruited all participants during household visits.

We tested two modes of data collection in a parallel randomized trial: face-to-face (FTF) interviewing and audio computer assisted interviewing (ACASI). FTF is the standard approach in household surveys: an interviewer asks questions and records respondents’ answers. In ACASI, respondents are equipped with headsets through which they hear pre-recorded questions; then they enter answers on their own, using a keypad.^37^ We hypothesized that ACASI might improve reporting of the HIV status of deceased siblings because it is more confidential than FTF.^38^

Participants were randomized 1:1 to FTF or ACASI. We stratified the randomization by participant gender and mortality of their known siblings (i.e., whether they had a deceased sibling who was a PHIV). All interviews were conducted in the local language (Chitumbuka). Interviewers did not have access to reference data on family composition, HIV status and mortality. For the ACASI version of the questionnaire, a team member pre-recorded questions and instructions in Chitumbuka. We designed images indicating where respondents should push on the screen if they wanted to enter a specific answer. We used color schemes with high contrasts and symbols to ensure that color-blind participants could use ACASI. ACASI included a training section, wherein we asked respondents simple questions so they could become familiar with self-interviewing. Respondents assigned to ACASI switched to FTF if they reported vision problems, e.g., not being able to see the screen in order to answer questions. To prevent selective manipulation of study group assignments, interviewers learned the allocation of respondents to ACASI or FTF only after an individual had consented to participate and his/her vision had been assessed. All data were collected using Open Data Kit (ODK).

We used FTF to ask questions about the respondent’s background (e.g., age, educational level) and to collect the standard components of siblings’ survival histories (i.e., names, sex and age of siblings). When respondents assigned to ACASI reported adult deaths, interviewers handed them the tablet and headset to answer questions about that death (i.e., HIV questions, accidents/injuries, and pregnancy/childbirth). Respondents assigned to FTF were asked these questions directly by the interviewer. We collected metadata on the interview from ODK (e.g., time spent by question).

Following data collection, 2 reviewers used data on names and sex to independently link the lists of maternal siblings reported by survey respondents to HDSS records. For each match between lists, they attributed his/her HDSS ID to the reported sibling. When reviewers disagreed in their matching outcomes, a third reviewer adjudicated. To avoid confirmation bias, none of the reviewers had access to survey or reference data on siblings’ vital status, ages, or HIV status.

### Data analysis

We described the background of respondents by assigned study group, including ownership of a mobile phone and prior use of the Internet. We included these latter variables to assess respondents’ familiarity with digital tools similar to ACASI. We described the reported characteristics of siblings (sex, age). We assessed the proportion of reported adult deaths for which we could not ascertain HIV status from survey data and explored whether this proportion varied by reported time since death. We created a binary variable taking value 1 if the death had occurred within 8 years of the survey (“recent deaths”), and 0 otherwise (“earlier deaths”).

The pre-specified primary outcomes were the sensitivity and specificity of survey data on HIV status of deceased siblings. We defined sensitivity as the proportion of deaths to PHIV according to reference data that were also classified as such from survey data. Specificity was the proportion of deaths to persons who were HIV-negative according to reference data that were also classified as such from survey data. The amount of time spent answering HIV questions – measured from ODK metadata– was a secondary outcome of the trial. All study group comparisons were conducted with participants included in their assigned study group (intent-to-treat). All analyses of study outcomes used survey weights to account for differences in selection probabilities and participation rates between HDSS residents in different sampling strata. We assessed differences in proportions (e.g., sensitivity) between study groups using chi-square tests, and differences in interviewing times using non-parametric rank sum tests.

Partial verification bias might affect analyses of primary outcomes,^39^ because reference data on the HIV status of deceased siblings are only available for a subset of the deaths reported during survey interviews. This usually leads to sensitivity estimates that are too high and specificity estimates that are too low.^40^ We imputed missing reference data on HIV status as recommended.^40,41^ We assumed that reference data were missing at random (MAR), i.e., their availability depended on reported characteristics of the deceased. We included sex, age at death, time since death and survey-assessed HIV status of the deceased in imputation models. We created 20 imputed datasets through chained equations, and we combined results using standard techniques.^42,43^ We did not pre-specify these adjusted analyses.

## Results

The study was conducted between October 12^th^, 2018 and January 14^th^, 2019. We allocated 307 HDSS residents to FTF and 306 to ACASI. Participation rates were 88.6% in the FTF group (272/307) and 87.3% in the ACASI group (267/306).

**Figure 1:**
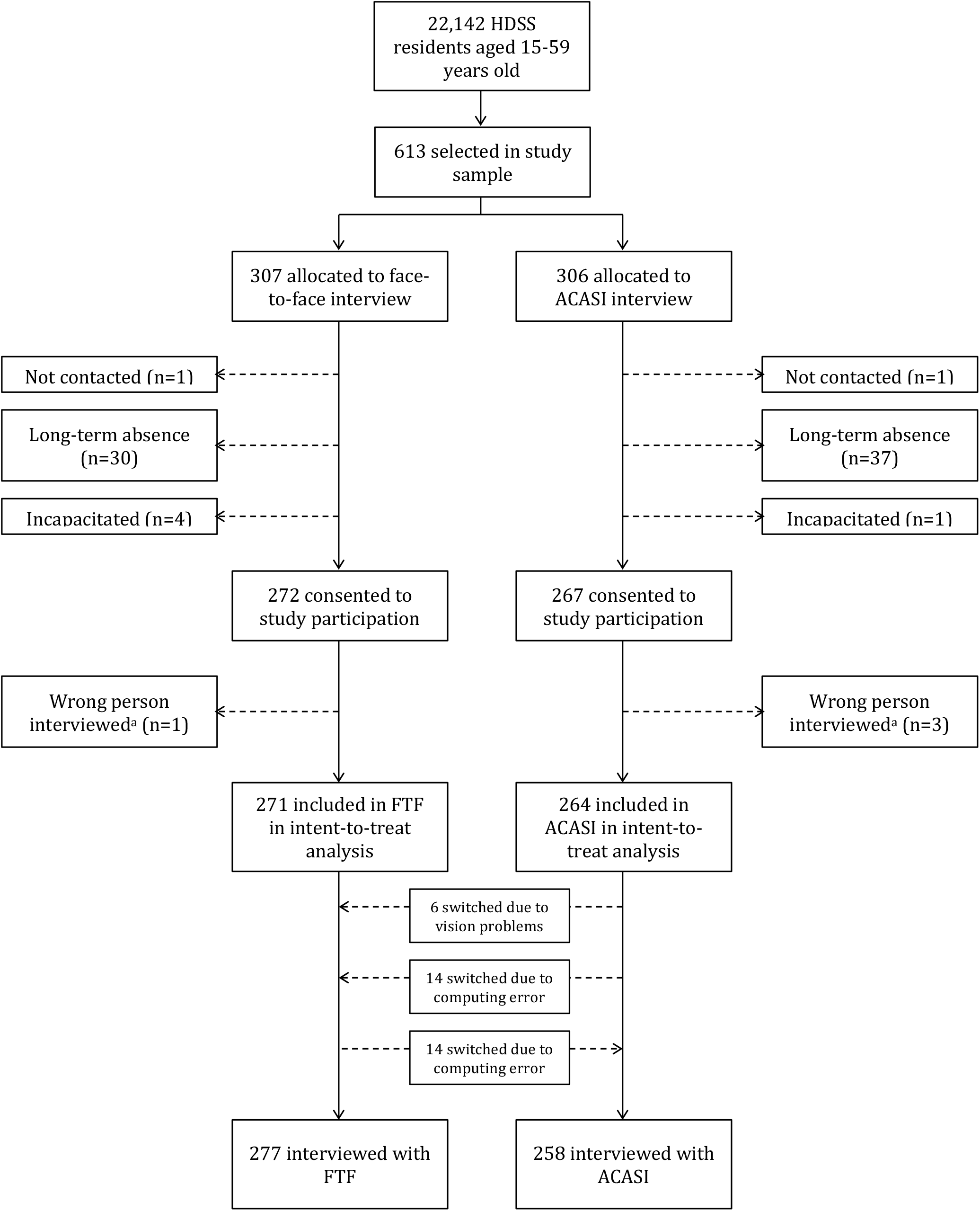
Enrollment process

*Notes: ^a^* These errors were first detected at the data editing stage, due to large discrepancies in age of the respondent between survey and HDSS data (>20 years); they were subsequently confirmed during household re-visits by study supervisors.

The educational level of participants was low (table 1), with more than a third of respondents not having completed primary education. Few respondents had prior experience of digital tools, e.g., slightly more than 1 in 10 had ever used the internet. The number of known siblings based on HDSS data varied across respondents: approximately 5% of respondents had no known sibling, whereas 48.5% of respondents in the ACASI group and 44.3% in the FTF group had 4 or more known siblings. Respondents listed 3,414 maternal siblings during survey interviews. There were 885 reported adult deaths: 427 in the ACASI group and 458 in the FTF group. In the FTF group,38.2% of reported deaths were recent vs 44.6% in the ACASI group. The complete reported distributions of ages at death, and dates of death, are available in appendix A2.

**Table 1:**
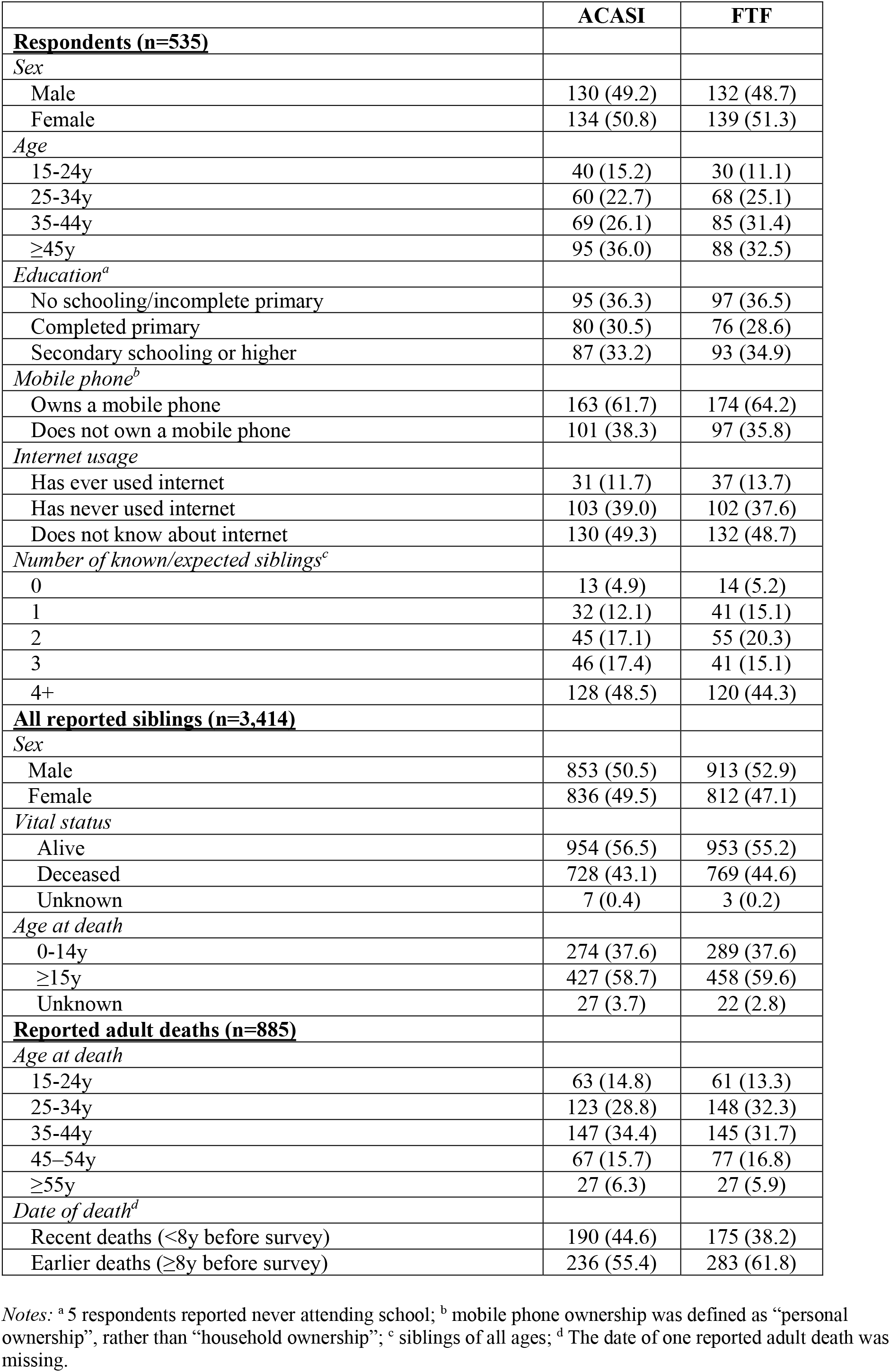
Characteristics of study participants, by assigned study group.

Based on data generated by the HIV questions (appendix A3), the HIV status of the deceased sibling could not be determined for 6.8% of adult deaths reported by ACASI respondents (95% CI = 4.4 to 9.2) and 13.5% of deaths reported by FTF respondents (95% CI = 10.4 to 16.7, figure 2). Among recent deaths, missing survey data on HIV status was as common in ACASI (5.3%, 95% CI = 2.1 to 8.4) and FTF (6.3%, 95% CI = 2.7 to 9.9). Among earlier deaths, the proportion of siblings whose HIV status could not be ascertained from survey data was lower in ACASI data(8.0%, 95% CI = 4.6 to 11.5) than in FTF data (18.0%, 95% CI = 13.5 to 22.5).

**Figure 2:**
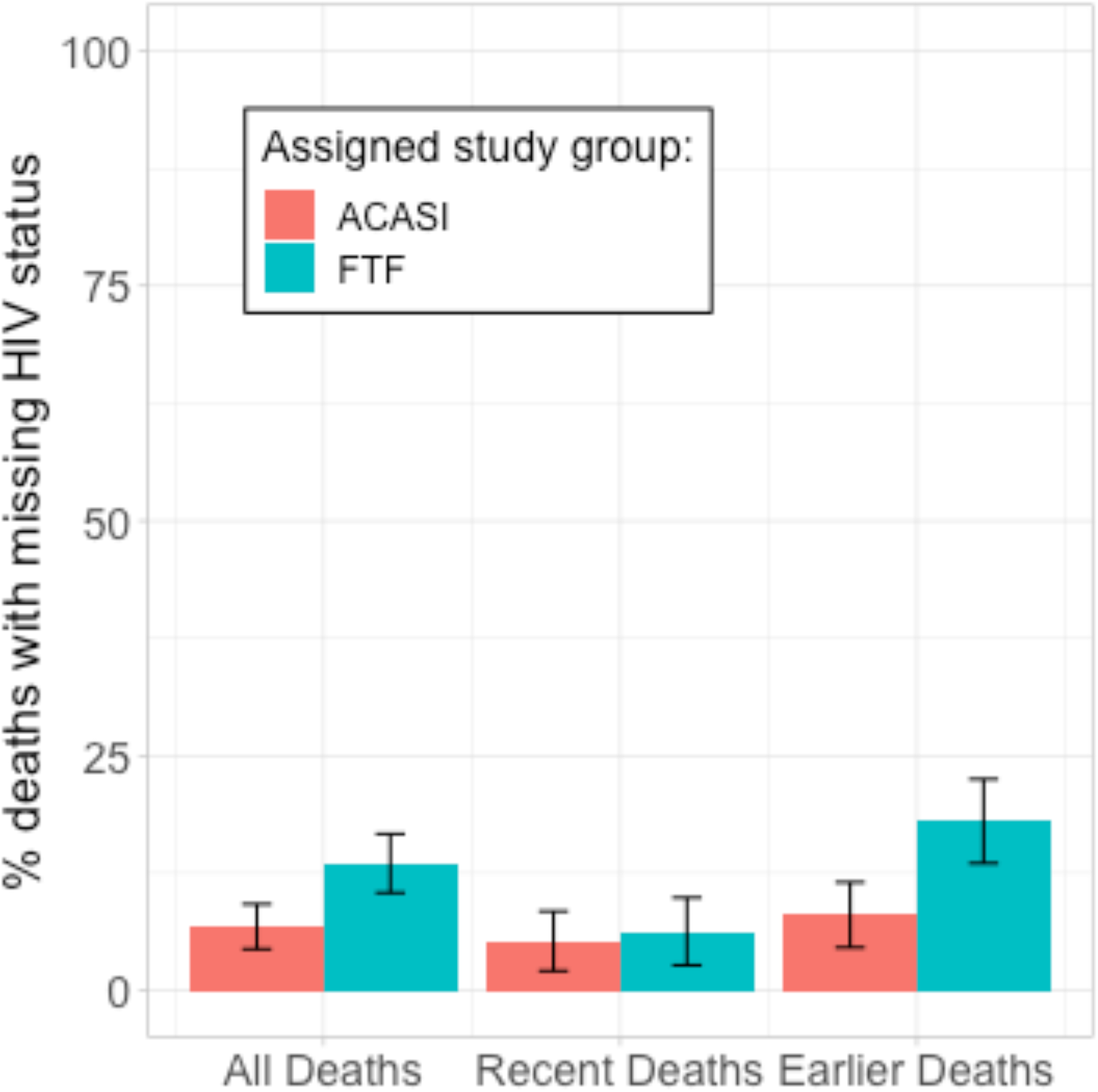
percentage of deceased adult siblings with missing data on HIV status, by assigned mode of interview and reported timing of the death (n = 885)

*Notes:* Recent deaths are deaths that have occurred over the past 8 years according to reports from survey respondents. Earlier deaths are deaths that have occurred more than 8 years ago according to reports from survey respondents. Vertical bars represent 95% confidence intervals. The calculations of these confidence intervals accounted for the clustering of adult deaths within families/respondents, and for the stratified survey design. All calculations used the “survey” package in R to incorporate survey weights. One respondent did not report the time since the death of her adult sibling; as a result, the analyses by reported timing of the death were conducted with n = 884.

Reference data on HIV status of the deceased were available for 262 of 885 reported adult deaths (29.6%, appendix A4). Among those, 180 deceased siblings were PHIV, and 82 were HIV-negative individuals, according to reference data. The availability of reference data on HIV status did not vary between study groups. However, reference data were more frequently available for deaths that were classified as deaths of PHIV on the basis of survey data.

Among reported deaths with reference data on HIV status, sensitivity was 0.89 (95% CI = 0.85 to 0.92, table 2) in the ACASI group and 0.91 (95% CI = 0.88 to 0.93) in the FTF group. After accounting for partial verification bias, estimates of sensitivity declined to 0.82 (95% CI = 0.46 to 0.96) in the ACASI group and 0.78 (95% CI = 0.44 to 0.94) in the FTF group.

**Table 2:** sensitivity/specificity of survey data in recording HIV status of deceased adult siblings, by assigned mode of interview.

|  | ACASI | FTF | P-value |
| --- | --- | --- | --- |
| **<u>Sensitivity</u>** |  |  |  |
| *Reference data*<sup>a</sup> | 0.89 (0.85 to 0.92) | 0.91 (0.88 to 0.93) | 0.40 |
| *Imputed data*<sup>b</sup> | 0.82 (0.46 to 0.96) | 0.78 (0.44 to 0.94) | 0.28 |
| **<u>Specificity</u>** |  |  |  |
| *Reference data*<sup>a,c</sup> | 0.94 (0.90 to 0.97) | 0.91 (0.86 to 0.95) | 0.71 |
| *Imputed data*<sup>b</sup> | 0.98 (0.79 to 1.00) | 0.96 (0.81 to 0.99) | 0.39 |

Notes: <sup>a</sup> Reference HDSS data were available for 262 deaths reported during the survey, 180 of which were deaths of persons who were HIV-positive and 82 of which were deaths of persons classified as HIV-negative.
<sup>b</sup> We imputed the reference HIV status when survey data on HIV status were not missing (n=794). We generated 20 datasets through chained equations, in which we imputed missing values of reference HIV status. We did so using the “mice” package in R.
<sup>c</sup> The null hypothesis of a one-sided test of the non-inferiority of ACASI relative to FTF with $\delta = 0.08$ was rejected with $p = 0.029$ . In this test, the null hypothesis assumes that the specificity of ACASI is inferior to the specificity of FTF by $\delta = 0.08$ .

In the subset of deaths for which reference data were available, the specificity of survey data on HIV status of the deceased was 0.94 (95% CI = 0.90 to 0.97) in ACASI and 0.91 (95% CI = 0.86 to 0.95) in FTF. Estimates of specificity increased when we accounted for partial verification bias (0.98 in ACASI vs. 0.96 in FTF).

ACASI training lasted a median time of 7.4 minutes (appendix A5). Asking the HIV questions required a median time of 0.4 minutes per deceased adult sibling in the FTF group and 0.7 minutes in the ACASI group (figure 3). In FTF interviews, this is comparable to the median time required to collect information on pregnancy-related deaths (0.4 minutes in FTF) and on accidental/external deaths (0.2 minute in FTF).

**Figure 3:**
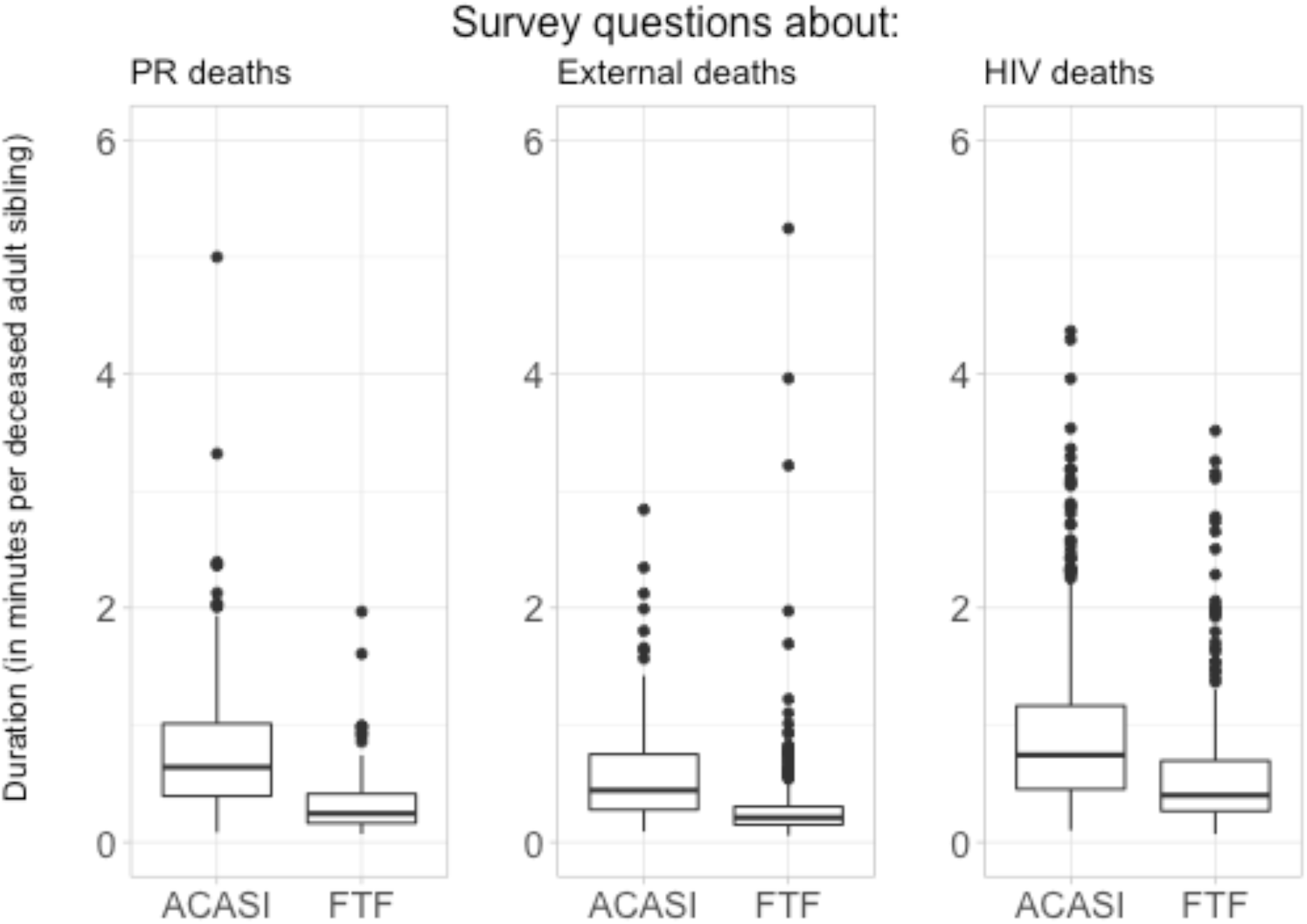
Distribution of time required to collect data on circumstances of reported adult deaths, by study group

*Notes*: PR = pregnancy-related; For each circumstance of death (i.e., PR deaths, external deaths and HIV deaths), based on rank sum tests, we rejected the null hypothesis that the distribution of interviewing time was similar in the ACASI and FTF arms (p< 0.001). All estimates reported in this figure were calculated per reported adult death (i.e., n = 885) for external deaths and HIV deaths; for PR deaths, these estimates were calculated for deaths of women of reproductive age (n = 384).

## Discussion

We tested whether accurate data on HIV-related mortality could be collected during household surveys. Such surveys already constitute the main source of representative data on all-cause and pregnancy-related mortality in most African countries. We thus added HIV questions to the standard adult and maternal mortality of the DHS questionnaire and conducted a validation study of these data in Karonga district in northern Malawi.

Adding HIV questions resulted in limited amounts of missing data on the HIV status of respondents’ deceased siblings, particularly among deaths that had occurred within 8 years of the survey (5–6%). This is important because adult mortality rates are often estimated solely from survey data on recent deaths and person-years.^45^

The added HIV questions generated a classification of deceased siblings according to their HIV status that was often accurate. In analyses accounting for partial verification bias, approximately 8 out of 10 deceased siblings who were PHIV were correctly reported as such by survey respondents. Our estimates of the specificity of survey data on HIV status were 0.96–0.98 in adjusted analyses. These levels are comparable to estimates of the sensitivity/specificity of survey data in recording pregnancy-related and injury-related deaths.^46–51^ Whereas questions about these latter circumstances are included in most recent DHS and MICS, HIV questions have not yet been included. This is a missed opportunity because collecting additional HIV-related data requires limited time: in face-to-face interviews, asking HIV questions took 0.4 minute (i.e., 25 seconds) per deceased adult sibling, on average.

We hypothesized that ACASI might yield more accurate data than FTF, but we found limited support for this hypothesis. ACASI generated less missing data than FTF, but only for deaths that occurred more than 8 years prior to the study. ACASI data did not have higher sensitivity than FTF data, but it required more time to collect mortality data. Due to ACASI’s limited effectiveness in improving accuracy, and the high burden it places on respondents, our study does not justify recommending the use of ACASI to collect mortality data in household surveys.

There are several limitations. First, our reference data do not constitute a gold standard in ascertaining the HIV status of deceased siblings. They rely partly on linkage with clinical registers from health facilities to establish the HIV status of the deceased. These linkages might be made erroneously, e.g., in the case of two persons with similar names. This might lead to estimates of the sensitivity/specificity of survey data that are too low. Reference data on HIV status were occasionally based on self-reports (appendix A4). Since self-reported data on HIV test results might be affected by social desirability bias, our reference classification might include PHIVs who were incorrectly classified as being HIV-negative.^22^ This would lead to estimates of the specificity of survey data that are too low. We also classified deceased siblings as HIV-negative if they had received negative test results within 5 years of their death. This definition might miss individuals who have seroconverted within a few months/years prior to death. Given low (recent) levels of HIV incidence in Malawi^52^ and patterns of disease progression however, such misclassifications are likely rare in our reference data.

Second, reference data on HIV status were only available for a subset of the deaths reported during the survey interview. We addressed this partial verification bias through multiple imputations of the reference data. Multiple imputations assumed that reference data were missing at random (MAR) among reported deaths. If the availability of reference data was also determined by other unobserved factors, then reference data were missing not at random and multiple imputations might not have adequately adjusted our estimates of sensitivity/specificity.

Third, the results of our study might not reflect the levels of sensitivity/specificity that could be achieved if HIV questions were included in household surveys such as DHS. This is so because in Karonga, the scale-up of HIV-related services has occurred earlier than in similar settings. Between 2005 and 2011, the HDSS population was targeted by multiple HIV serosurveys, during which adult residents were offered home-based HIV testing. A large fraction of PHIV in the area became aware of their HIV status and might subsequently have disclosed this information to their siblings.^53^ In other settings where HIV testing was not scaled-up in a similar manner, the accuracy of survey data in recording the HIV status of deceased siblings might be lower.

Fourth, study participants were residents of the HDSS area, whose siblings had died when they also resided in this area. Due to this geographic proximity, our study might have included respondents who had increased knowledge of the HIV status of their deceased siblings. The accuracy of survey data on HIV status of deceased siblings might be lower among family members who resided further away from the deceased. However, this might not necessarily be the case since migrants are often central in providing the financial support required to care for PHIV. They might also return home to visit sick relatives and might otherwise remain in touch with their siblings through mobile communications.

Finally, due to limited sample size, we did not conduct sub-group analyses. We thus did not test whether the sensitivity/specificity of survey data on HIV status of deceased siblings might vary with characteristics of a) the respondents (e.g., gender, educational levels), b) the deceased siblings (e.g., gender, age at death, time since death), or c) the interviewer. We also did not investigate whether ACASI might help improve the accuracy of survey data for some population groups, e.g., those with secondary schooling and/or prior experience with digital tools.

In conclusion, we suggest a simple strategy for improving the measurement HIV/AIDS mortality in African countries with limited data. Adding a few questions to questionnaires widely used by household surveys might generate data on the excess mortality that persons with HIV experience in a population. Such questions require limited additional time to collect, but they might help calibrate the statistical models used to track progress towards global HIV elimination objectives.

## Data Availability

All study data are available upon requests, and will be made available shortly after preparation of a repository.

## Appendix A1 mathematical framework

We outline a framework detailing how the data collected through the HIV questions that we added to siblings’ survival histories might help better assess the contribution of HIV to adult mortality in affected SSA countries.

We note *S* the number of siblings (including selected respondents) listed during a survey that included a sibling survival history module (e.g., a DHS). We note *D* a binary variable indicating that one of these siblings died during a given timeframe, from any cause of death. We note *H* a binary variable that indicates whether a sibling has been infected with HIV. For simplicity, we consider that all deaths occur exactly at the same time (e.g., during a reference period). This assumption can easily be relaxed in future refinements of this framework, given that data on the timing of deaths are collected during siblings’ survival histories.

The addition of the HIV questions to siblings’ survival histories then allows estimating 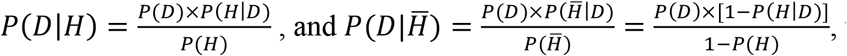, i.e., the proportion of deaths that occurred to PHIV. From Bayes’ rule, we have: 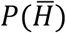, where *P*(*H*) is the proportion of all siblings who have become infected with HIV irrespective of survival status, and 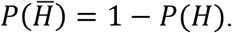 is the proportion of all siblings that have not become infected with HIV. Since *H* is a binary variable, we have 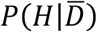. The mortality ratio associated with HIV infection, i.e., 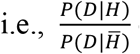 can thus be re-written as:

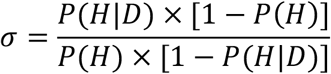

When σ > 1,PHIV die at faster rate than individuals who are HIV-negative, and σ measures the excess mortality faced by PHIV in the population of interest. Success in prevention, care and treatment of HIV can then be measured by the pace at which σ converges to 1 over time (e.g., from one survey to the next).

From the formula of total probabilities, we can expand *P*(*H*) and rewrite σ as follows:

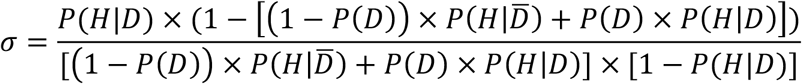

In our expression of σ, *P*(*H*|*D*) can be estimated from survey data, if the 3 HIV questions yield accurate data. Estimates of *P*(*H*|*D*) can be adjusted for potential misclassifications and other reporting errors, using data on sensitivity and specificity from validation studies such as the one reported in this paper.

*P(D)* can be estimated directly from the siblings’ survival histories. On the other hand, data on 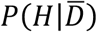, the prevalence of HIV among surviving siblings, are available through several modalities. In some surveys, 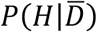, can be measured directly, through sero-testing of survey respondents (e.g., some DHS). In other surveys, 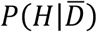 can also be measured through self-reported data, by asking respondents to report their most recent test results. These approaches require the assumption that the sample of survey respondents is a random sample of surviving siblings. In addition, data on 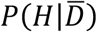 can also be potentially collected during siblings’ survival histories, by extending the HIV-related questions to all siblings, whether they are alive or deceased at the time of the survey.

## Appendix A2 Distributions of ages at death and time since death

**Figure A2-1:**
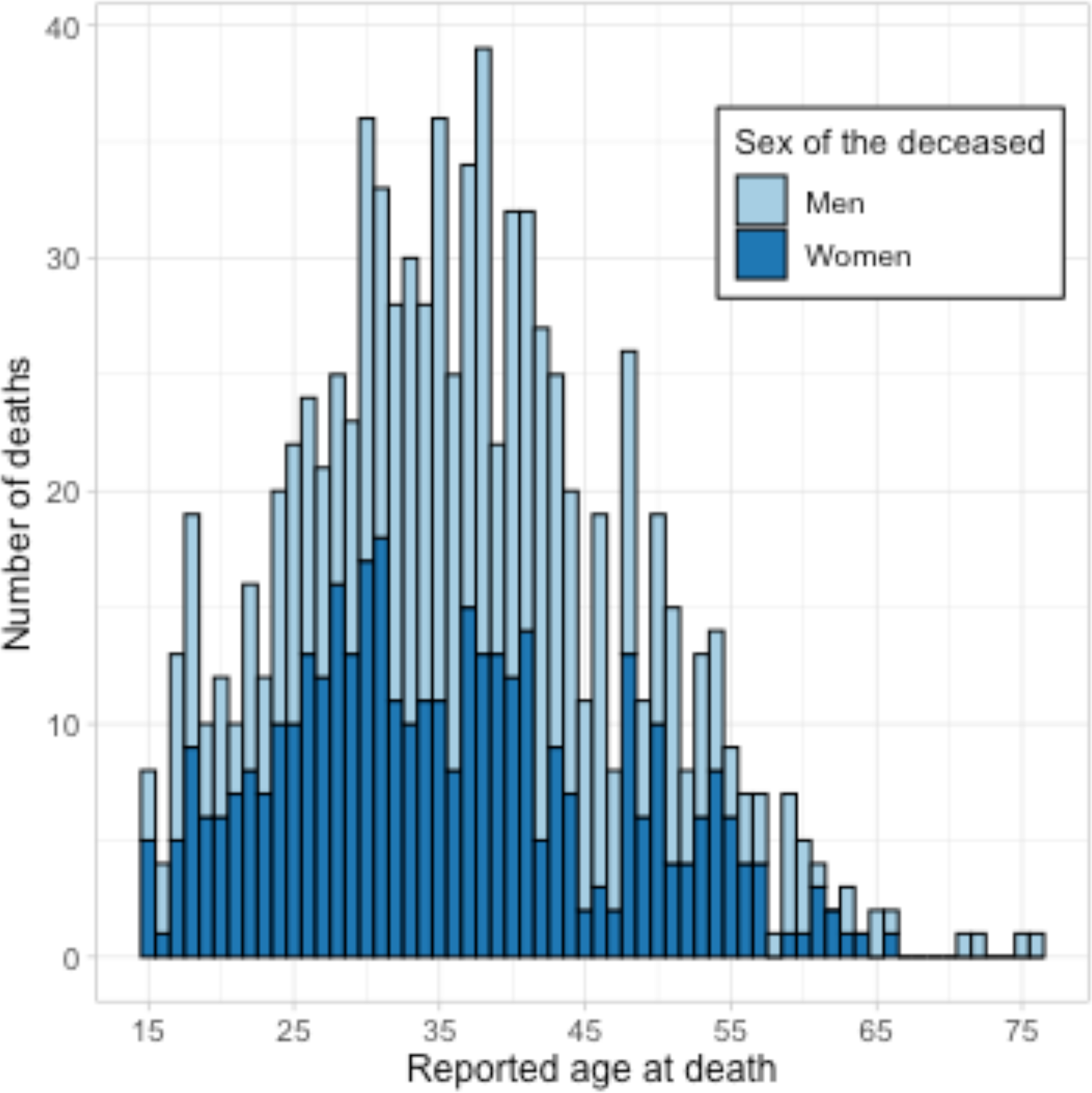
Reported ages at death, by sex of the deceased (n = 885)

**Figure A2-2:**
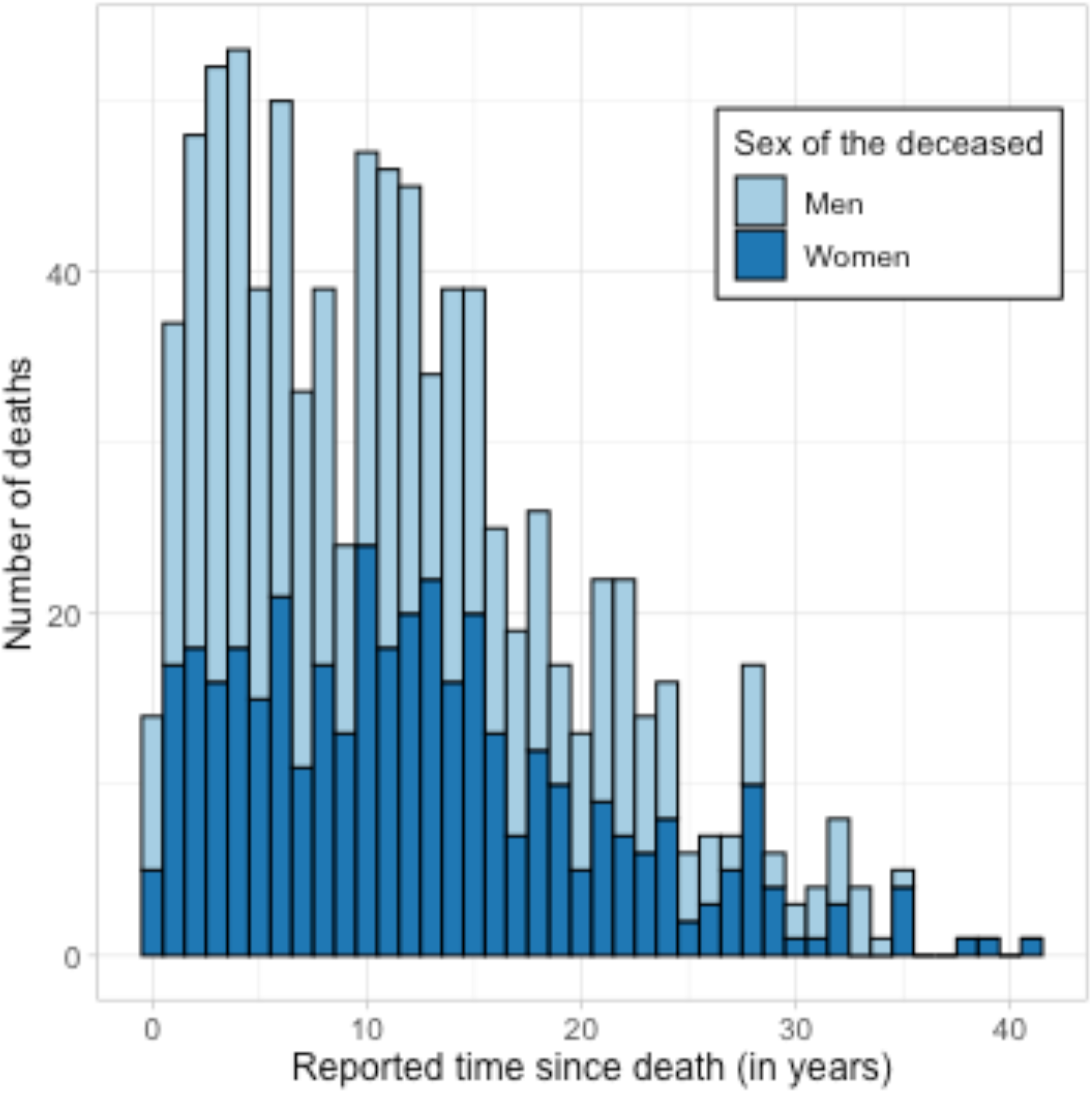
Reported numbers of years since the death, by sex of the deceased (n = 884).

*Notes:* the date of the death was not reported for one of the reported adult deaths.

## Appendix A3 data generated by HIV questions

**Figure A3-1:**
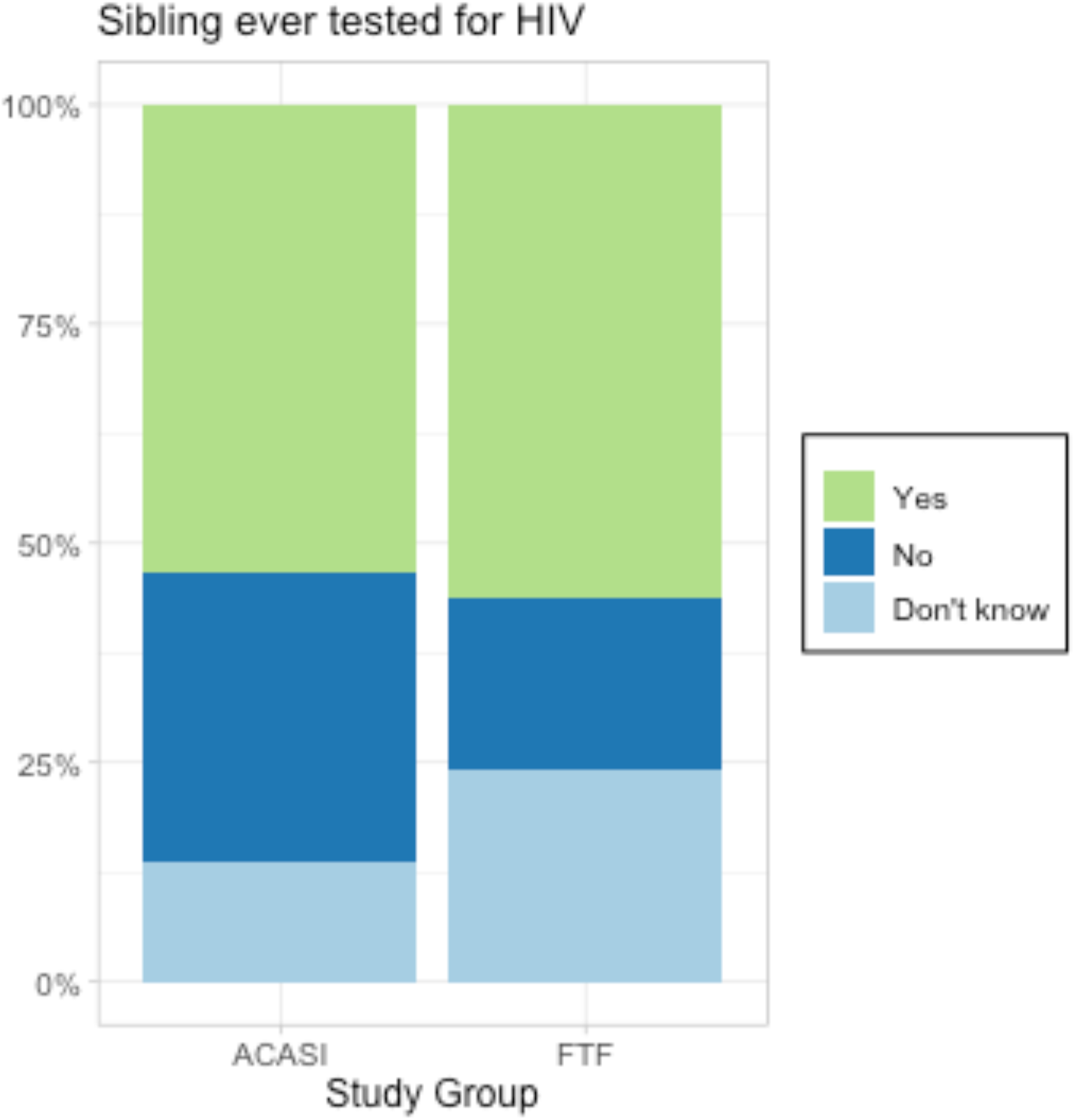
Distribution of answers to question “Has [deceased sibling] ever been tested for HIV?”, by study group (n = 885)

**Figure A3-2:**
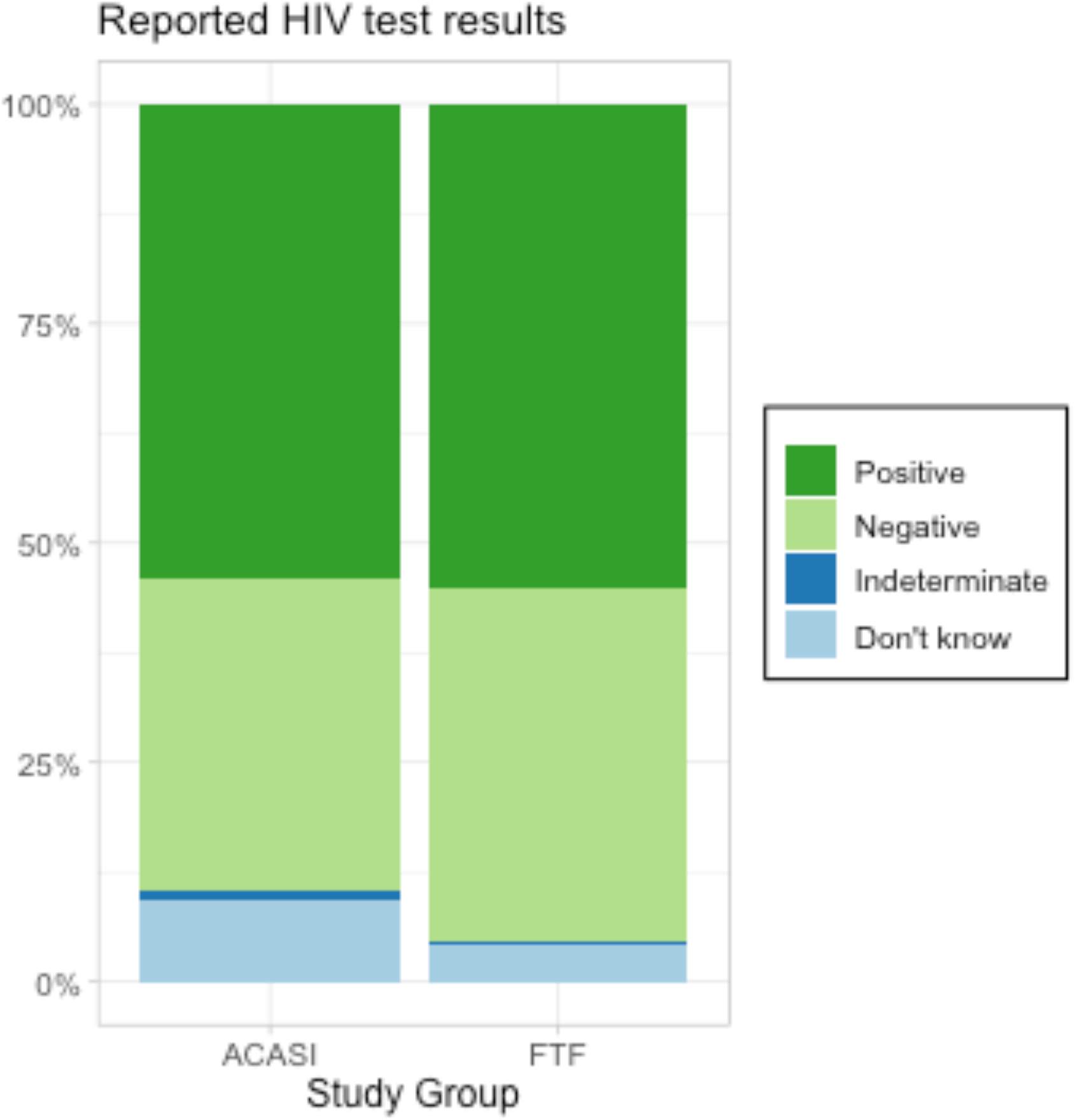
Distribution of answers to question “What were the results of [deceased sibling]’s latest HIV test”, by study group (n = 486)

**Figure A3-3:**
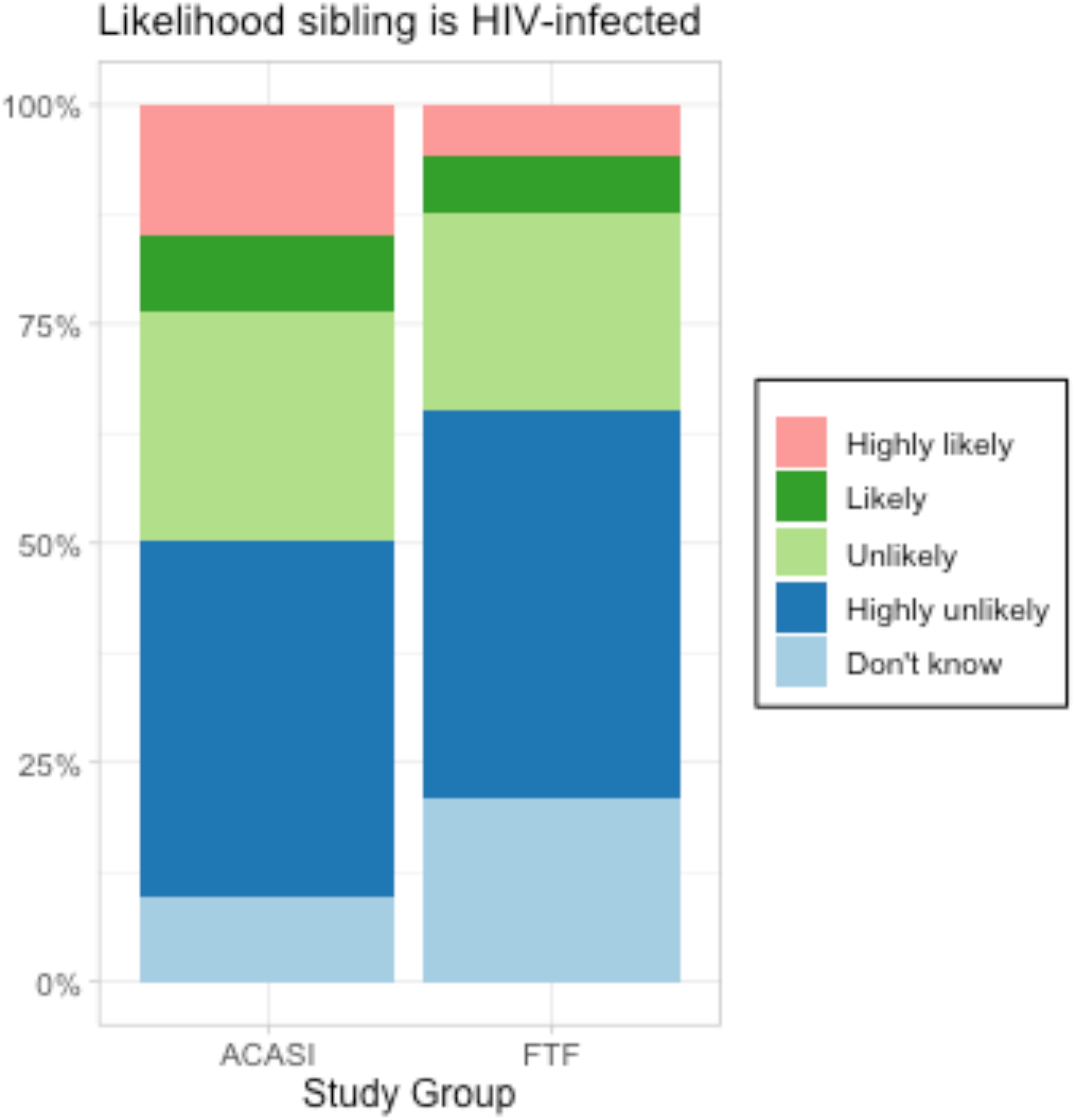
Distribution of answers to question “How likely do you think it is that [deceased sibling] was infected with HIV?”, by study group (n = 620)

## Appendix A4 Linkages between survey and reference data

**Figure A4.1:**
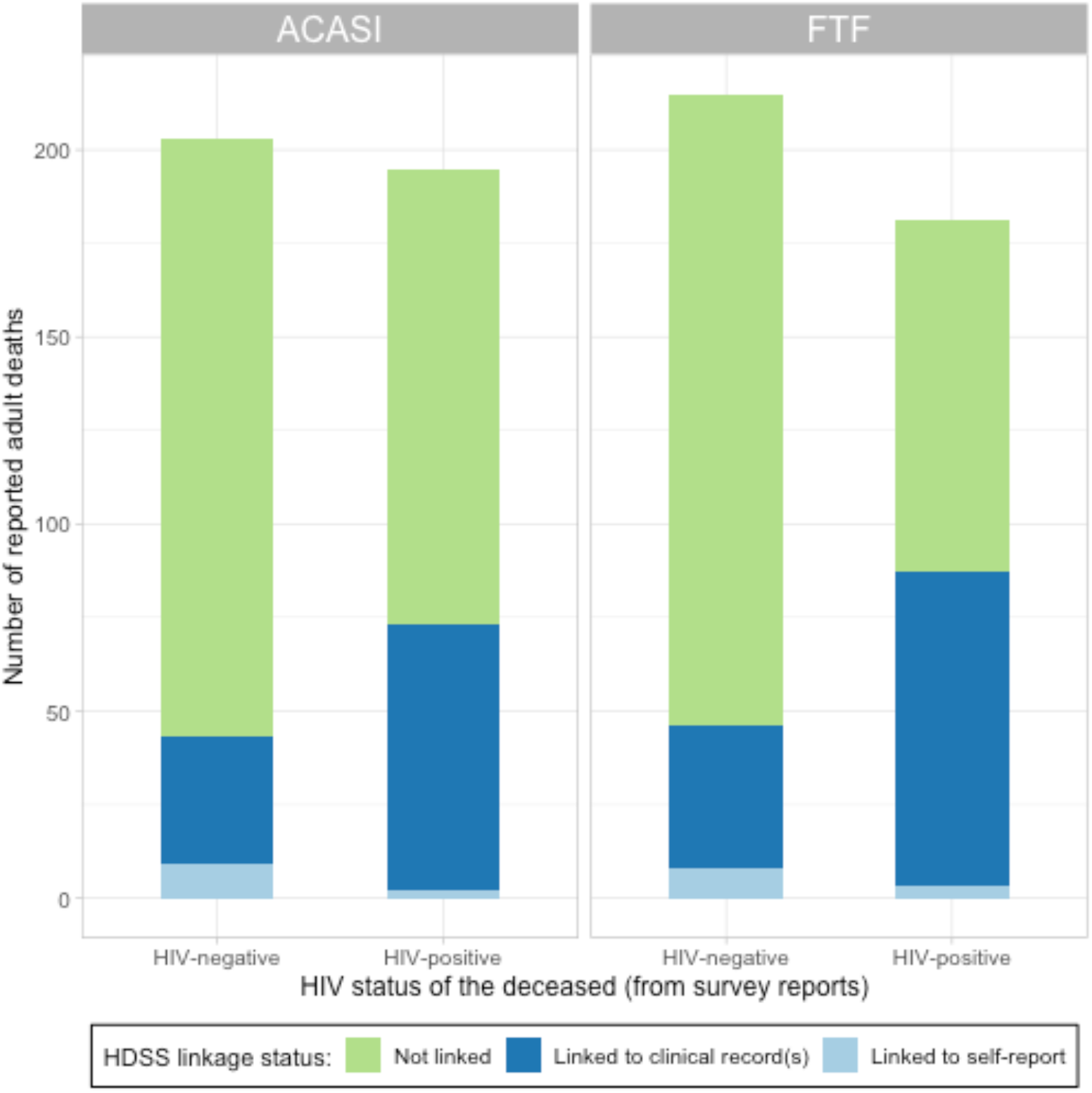
Outcomes of linkages between reported survey data and HDSS datasets, by reported HIV status of the deceased and study group.

*Notes:* Clinical records in the HDSS datasets include results of the serosurveys conducted between 2005 and 2011, as well as data routinely extracted from clinical registers at facilities serving the HDSS population.

**Figure A4.2:**
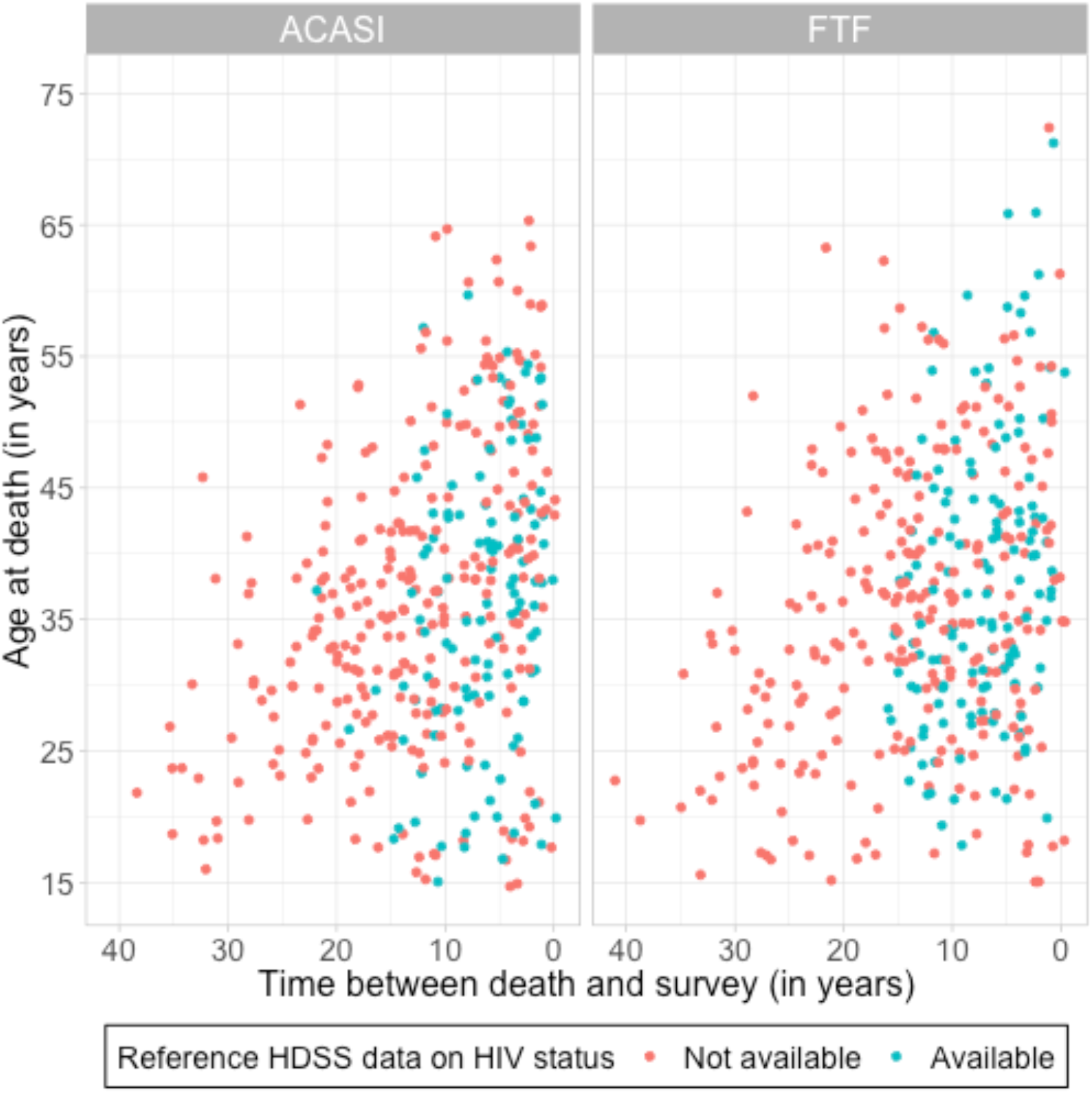
Joint distribution of reported ages at, and time since, death (n = 884), by study group and availability of reference HDSS data.

*Notes:* one death was not included on the graph due to missing data on date of death.

## Appendix A5 Duration of ACASI training

**Figure A5-1:**
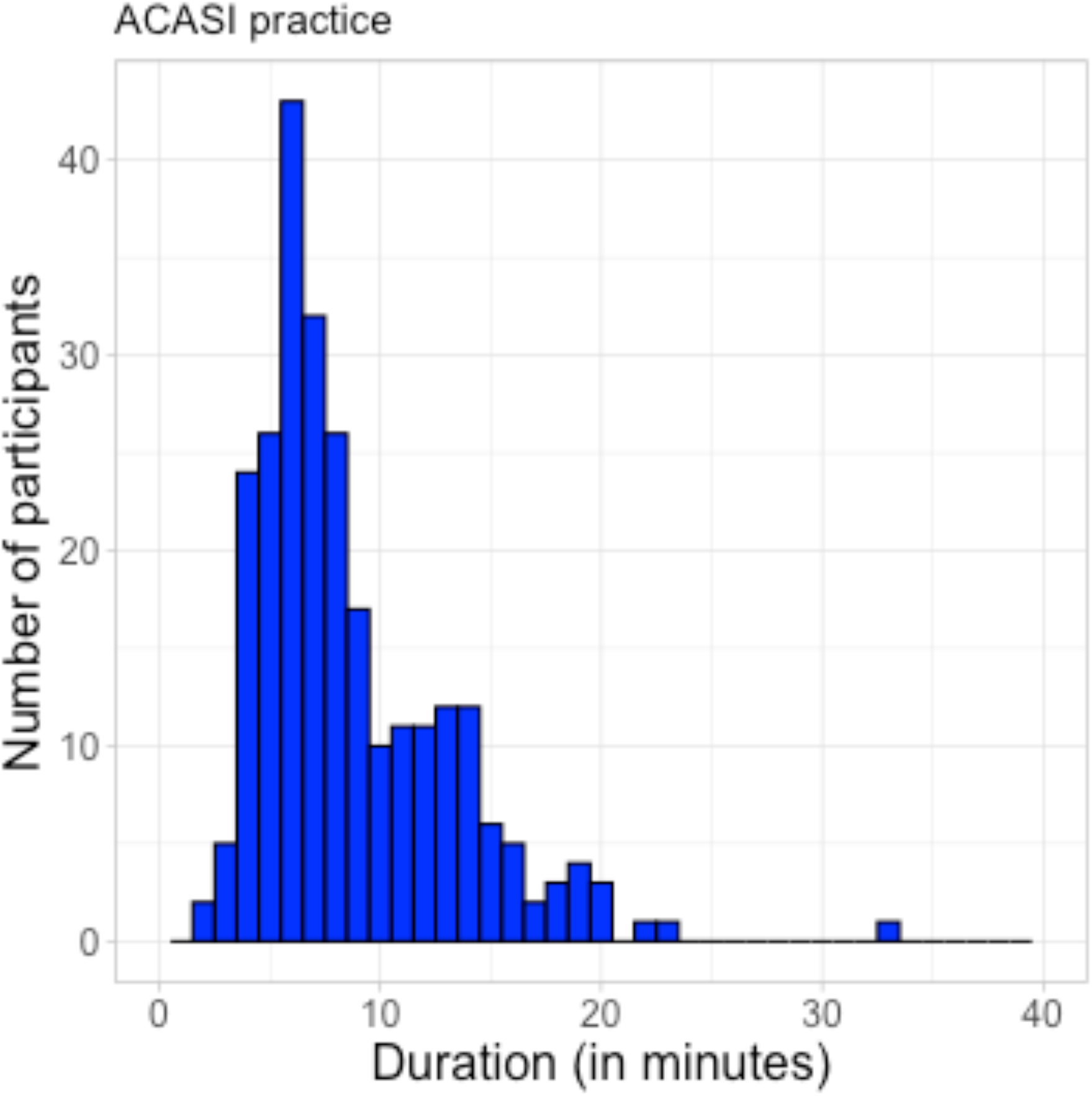
duration of ACASI practice.

Notes: duration estimates were obtained using the audit feature in ODK.

